# Oxymatrine Induces Autophagy and Apoptosis in Human Poorly Differentiated Gastric Adenocarcinoma Cells via Slit2/Robo1 Signals

**DOI:** 10.1101/2021.04.28.21256231

**Authors:** Mao-Tao He, Ting Zhang, Jing-ya Hei, Li-qiong Ma

## Abstract

Oxymatrine (OMT), is a natural quinoxaline alkaloid from the traditional Chinese medicine herb and has been shown to exhibit anticancer properties on various types of cancer cells. Poorly differentiated gastric adenocarcinoma is a common malignancy of gastric cancer that is more aggressive and has a poor prognosis. In the present study, we investigate the effects of Slit2/Robo1 signals in poorly differentiated gastric adenocarcinoma and adjacent tissues, and the anticancer properties of OMT on human poorly differentiated gastric adenocarcinoma BGC-823 cells and evaluate their underlying mechanisms. The expression levels of Slit2 and Robo1 proteins were measured in 20 pairs of human poorly differentiated gastric adenocarcinoma tissues and adjacent normal tissues using western blot. The expression of apoptosis related proteins and autophagy-related proteins was detected by western blot. The cells viability was detected by CCK-8 assay. The migration of BGC-823 cells was detected by transwell experiments. The expression of related proteins was detected by western blot. The result shows that Slit2 and Robo1 are significantly increased in poorly differentiated gastric adenocarcinoma. The apoptosis and autophagy are inhibited in poorly differentiated gastric adenocarcinoma. OMT inhibits the growth and migration of BGC-823 cells in vitro. OMT inhibits the activation of Slit2/Robo1 signals and induces apoptosis and autophagy in BGC-823 cells. These findings suggest that the antitumor effects of OMT may be the result of inhibition of cell growth and migration, and inhibits the activation of Slit2/Robo1 signals pathway and induces apoptosis and autophagy. OMT may represent a novel anticancer therapy for the treatment of poorly differentiated gastric adenocarcinoma.

## Introduction

Gastric cancer is one of the most common malignant tumors in humans, and its prognosis is related to the degree of tumor differentiation and metastasis^[1]^. Poorly differentiated gastric adenocarcinoma is a common malignancy of gastric cancer that is more aggressive and has a poor prognosis, mainly due to the biological characteristics of poorly differentiated gastric cancer, which makes it more prone to metastasis^[2]^. However, the current therapeutic effect and prognosis are still unsatisfactory, despite multimodal approaches involving surgery, chemotherapy, and radiotherapy. Thus, it is urgent to develop new anti-cancer agents or adjuvants for the treatment of patients with poorly differentiated gastric adenocarcinoma.

Oxymatrine (OMT) is a natural quinoxaline alkaloid and the primary biologically active ingredient extracted from the root of the Chinese herbal medicine Sophora flavescens Ait. OMT has a variety of pharmacological properties, including anti-inflammatory, anti-oxidative effects, anti-proliferative effects, and anti-apoptotic activities ^[3,4]^. Recent studies have found that OMT has good anticancer activities^[5]^. OMT has been shown to inhibit tumor cell proliferation in different cancer cells, including breast cancer cell lines (MCF-7), gastric cancer cells (SGC-7901 and MKN45), and human liver cancer cells (SMMC-7721). OMT induces cell cycle arrest and apoptosis via the PI3K/AKT/mTOR pathway in glioma^[6]^. Preliminary research showed the inhibitory effects of OMT on vascular and lymph node invasion in gastric cancer^[7]^. Thus, OMT may become a new therapeutic agent for the prevention and treatment of poorly differentiated gastric adenocarcinoma.

In recent years, studies have confirmed that Slit2/Robo1 signals play an important role in the occurrence and development of tumors. It has been reported that the slit2/robo1 pathway plays an important role in tumor angiogenesis^[8]^. Slit2 can promote the formation of new blood vessels in gastric cancer by acting on Robo1. Slit2/robo1 pathway promotes tumor growth and metastasis^[9]^. Studies have shown that miR-27a-3p/Slit2 can regulate apoptosis, autophagy, and inflammatory response in LPS-treated vascular endothelial cells^[10]^. Autophagy and apoptosis play important roles in tumor treatment. Some autophagy-related proteins such as Beclin-1 and LC3 have prognosis for gastric cancer. Autophagy and apoptosis are simultaneously involved in deciding the fate of cancer cells, where exists interaction between each other. However, the role of slit2/robo1 signals in gastric cancer and its possible mechanism are still unclear. Therefore, in this study, we explored the role of Slit2/Robo1 signals in the autophagy and apoptosis of poorly differentiated gastric adenocarcinoma during the treatment of OMT.

## Materials and methods

### Materials

The antibodies of Slit2, Robo1, Caspase3, Bax, Bcl2 and Beclin1 were purchased from Proteintech. Cleaved caspase3, and LC3B were obtained from Cell Signaling Technologies. Oxymatrine (B21470, HPLC ≥ 98%) used in this study was supplied by Shanghai Yuanye Bio-Technology Co., Ltd (Shanghai, China). β-Actin antibodies were purchased from Bioss (Beijing, China).

### Human cancer tissue samples

20 pairs of human poorly differentiated gastric adenocarcinoma tissues and adjacent normal tissues samples were obtained from the General Hospital of Ningxia Medical University from 2017 to 2019.Tissue specimens were surgically removed and pathologically diagnosed as poorly differentiated gastric adenocarcinoma after surgery. All patients didn’t receive radiotherapy and chemotherapy before surgery treatment, and didn’t accompanied by other malignant tumors. The study was approved by the ethics committee of General Hospital of Ningxia Medical University.

### Cell culture

Poorly differentiated gastric adenocarcinoma cell line (BGC-823) was cultured in RPMI-1640 medium (Hyclone), 10% fetal bovine serum (FBS), penicillin and streptomycin. Culture condition: 5% CO2, 37°C incubator.

### Cell viability assay

Cell viability was measured using Cell Counting Kit-8 (CCK-8) (Dojindo Molecular Tech. Inc., Rockville, MD) according to the manufacturer’s instruction. In brief, BGC-823 cells were plated into 96-well plates (10000/well, Corning Inc., New York, NY). After 24 h, the cultures exposed to various concentrations of oxymatrine (0, 2, 4, 6, 8 mg/mL). After 24 h of incubation, the results were read in the SpectraMax microplate reader (Molecular Devices, Sunnyvale, CA). The relative intensity of the untreated, control cells was arbitrarily converted to 100% cell viability, and experimental groups were converted to their corresponding percentages relative to the control.

### Transwell assay

BGC-823 cells (2×10^4^ cells/well) were seeded into the upper chamber of a polycarbonate transwell plate (8 *μ*m pores size; Corning, USA) in 300 *μ*L of a serumfree medium with OMT. 800 *μ*L of DMEM with 10% FBS was then added to the lower chamber as a chemoattractant. After 24 hours of culture, tumor cells in the upper chamber clustered in the lower chamber, and then the culture plate was removed from the incubator. A swab was used for removing the cells in the upper chamber. In addition, cells in the lower chamber were fixed in 4% paraformaldehyde and then 0.1% crystal violet was added for staining and then photographed and counted under high magnification.

### Western blot

Tissues or cells were lysed in RIPA lysis buffer containing protease inhibitor cocktails. Equal amounts of protein (50µg /well) were loaded into wells for 10% SDS-PAGE, and the resultant gel was transferred to PVDF membranes. The membrane was incubated with primary antibodies in blocking buffer at 4°C overnight. The following primary antibodies were used: Slit2 (1:1000), Robo1 (1:1000), caspase 1 (1:1000), cleaved caspase 1 (1:500), Bax (1:1000), Bcl2 (1:1000), LC3B (1:500), and Beclin1 (1:1000). β-actin (1:6000) were used as internal controls for protein loading. Then, the membrane was incubated with secondary antibodies at room temperature for 1 h. Imaging was performed using a AI600 imaging system, and the ratio of the target protein level to β-actin was calculated.

### Statistical method

Each experiment was independently repeated a minimum of three times. Statistical analysis was performed with IBM SPSS Statistics version 20.0. All values are expressed as the mean ± standard deviation (SD) or as a percentage of the control. The datasets were analyzed by ANOVA followed by the LSD post-hoc test. Statistical significance was set at *p* < 0.05.

## Results

### 1. Expression of Slit2 and Robo1 proteins in poorly differentiated gastric adenocarcinoma and adjacent tissues

In order to investigate the role of Slit2/Robo1 signals in gastric adenocarcinoma, the expression levels of Slit2 and Robo1 proteins were first measured in 20 pairs of human poorly differentiated gastric adenocarcinoma tissues and adjacent normal tissues using western blot. The results showed that the Slit2 and Robo1 expression levels were generally higher in the gastric adenocarcinoma tissues than in the matched normal gastric tissues (Fig. 1). This result showed that Slit2 and Robo1 were significantly increased in poorly differentiated gastric adenocarcinoma.

**Fig. 1.**
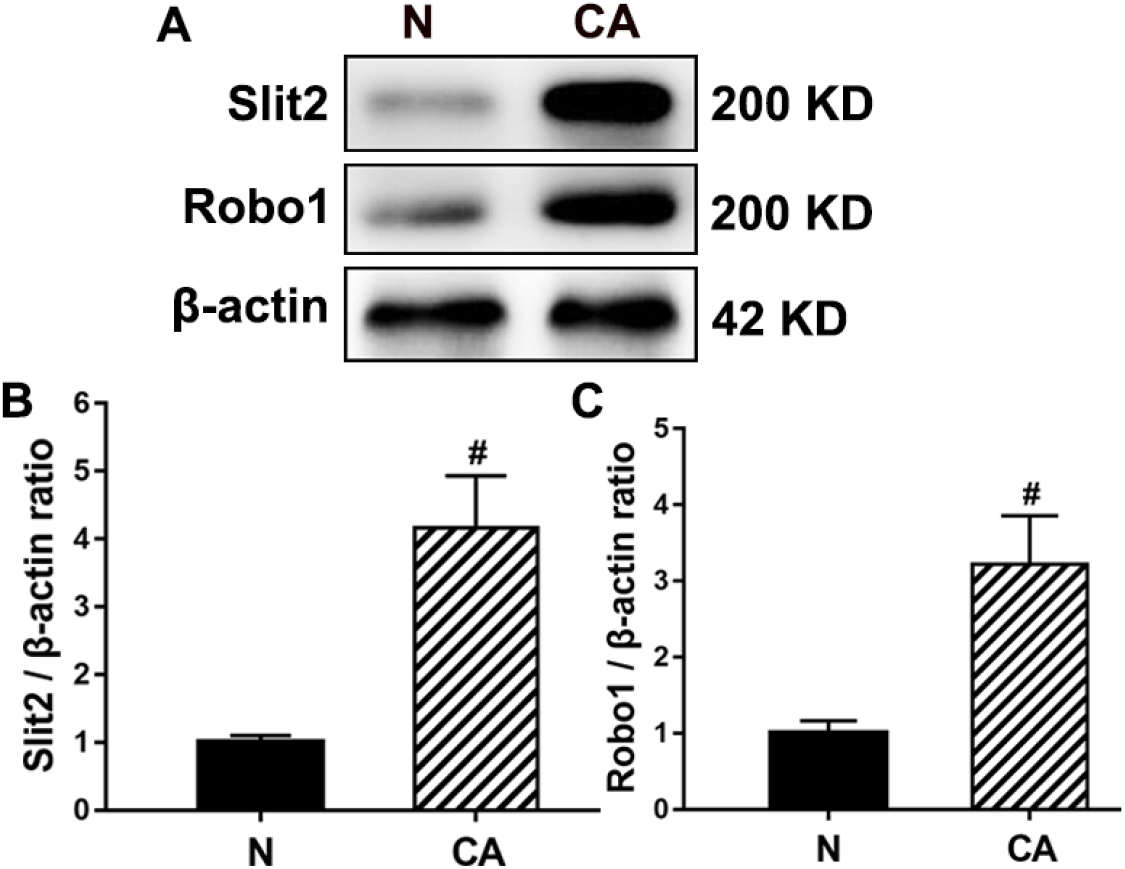
Expression of Slit2 and Robo1 protein in poorly differentiated gastric adenocarcinoma and adjacent tissues. (A) The expression of Slit2 and Robo1 were determined using western blot. (B) Image J densitometric analysis of Slit2/β-actin and Robo1/β-actin ratios from immunoblots.

**Fig. 2.**
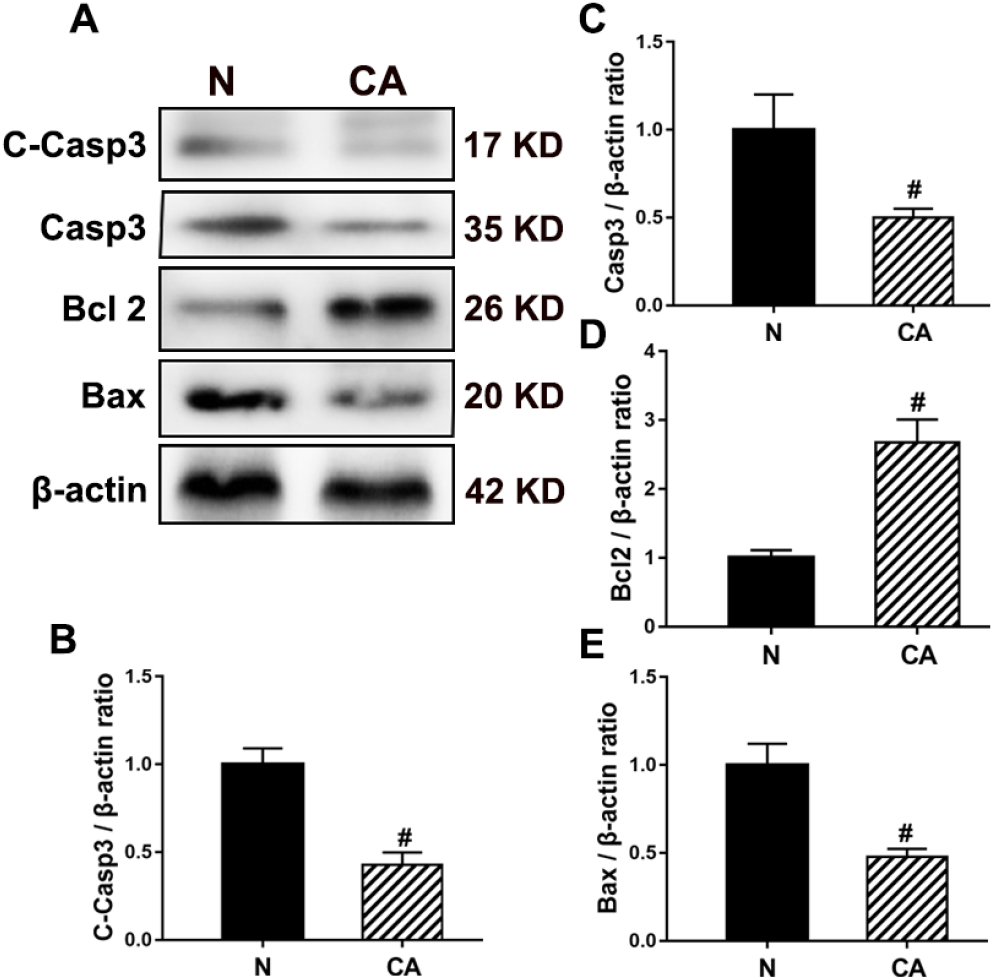
Expression of apoptosis related proteins in poorly differentiated gastric adenocarcinoma and adjacent tissues. (A) The expression of Cleaved caspase3, Caspase3, Bax, and Bcl2 were determined using western blot. (B-E) Quantification of Cleaved caspase3, Caspase3, Bax, and Bcl2 immunoblots.

**Fig. 3.**
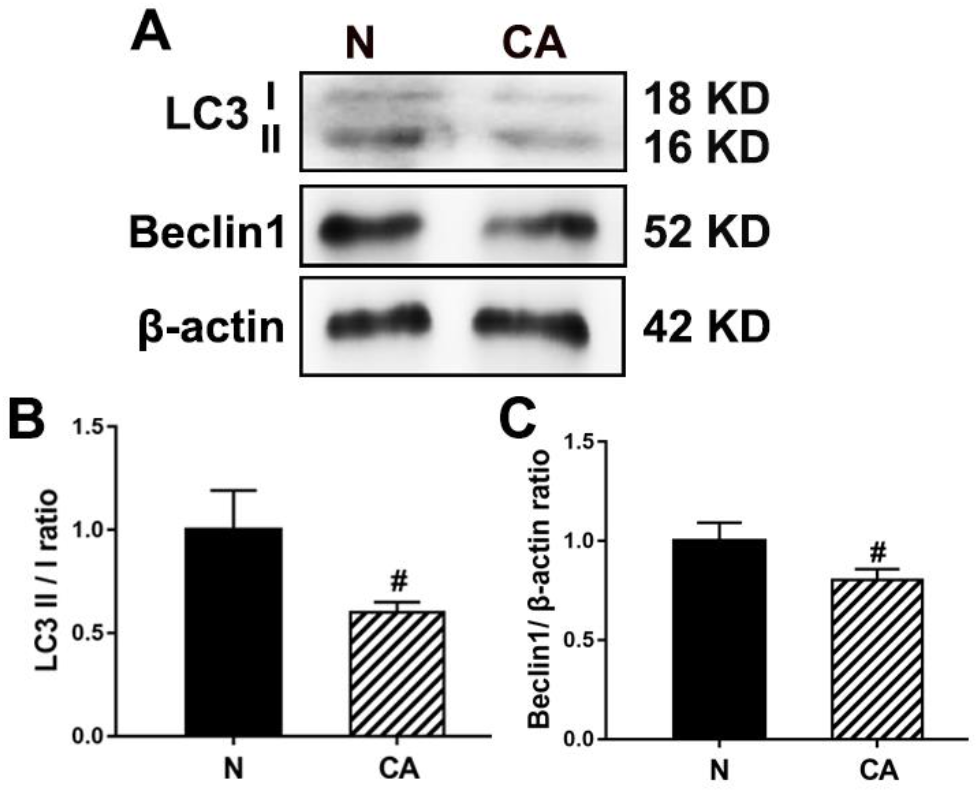
Expression of autophagy-related proteins in poorly differentiated gastric adenocarcinoma and adjacent tissues. (A) The expression of LC3 II/I and Beclin1 were determined using western blot. (B, C) Image J densitometric analysis of LC3 II/LC3 I and Beclin1/β-actin ratios from immunoblots.

**Fig. 4.**
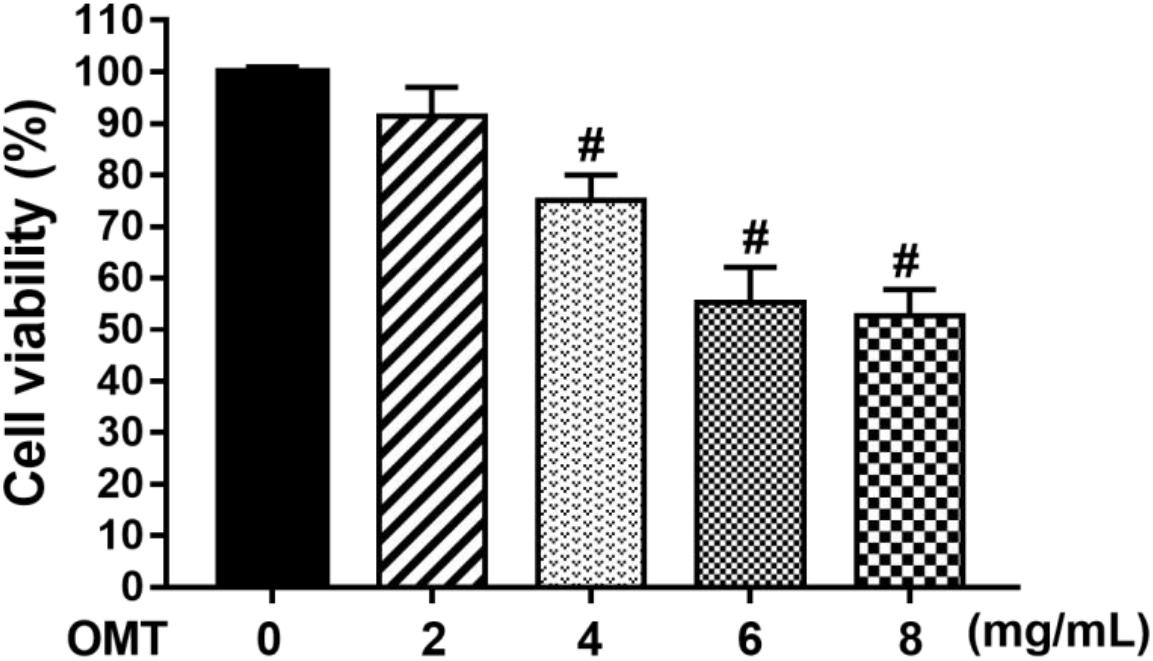
Oxymatrine inhibits the growth of BGC-823 cells in vitro. The cell viability of each group was determined by CCK-8 assay.

**Fig. 5.**
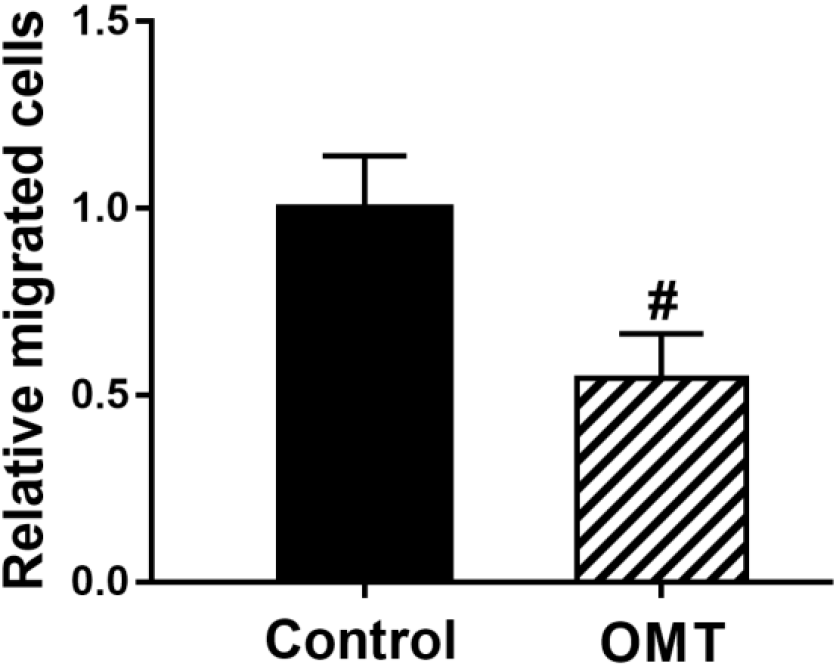
Oxymatrine inhibits the migration of BGC-823 cells in vitro. The cell viability of each group was determined by CCK-8 assay.

**Fig. 6.**
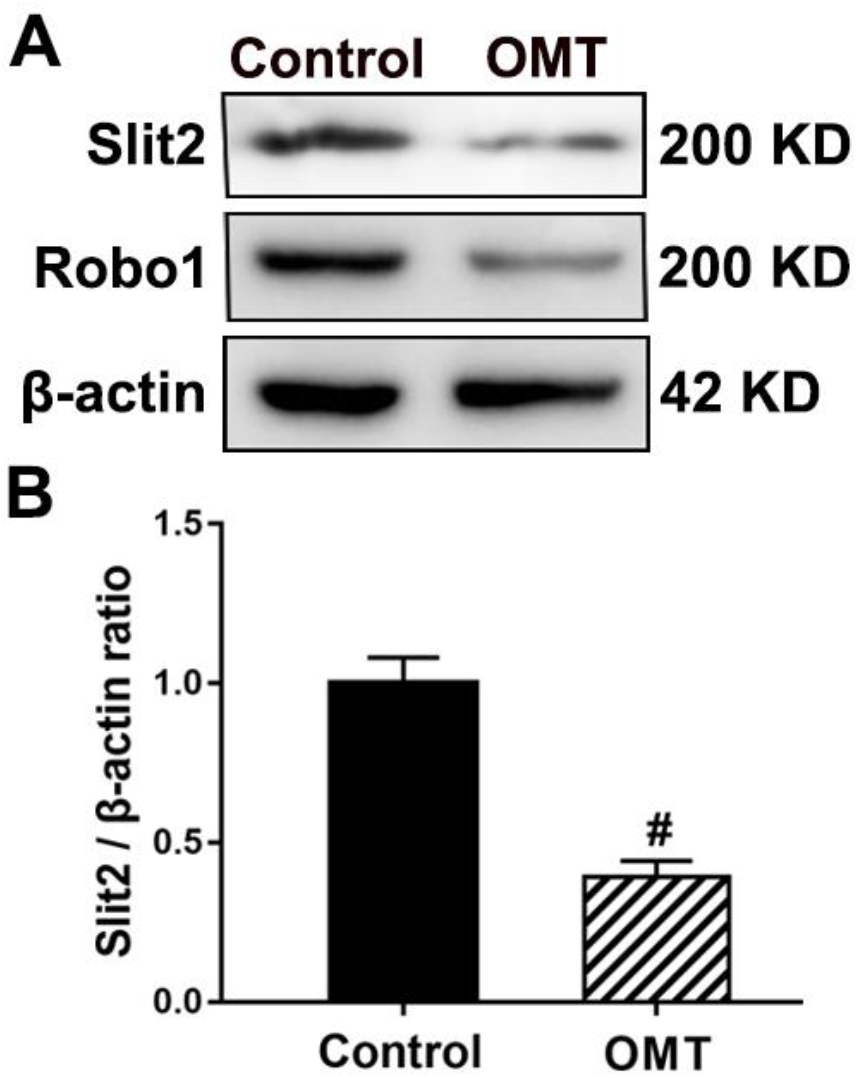
The effect of oxymatrine on the Slit2/Robo1 signals in BGC-823 cells. (A) Western blot results. (B) Western blot quantification histogram.

**Fig. 7.**
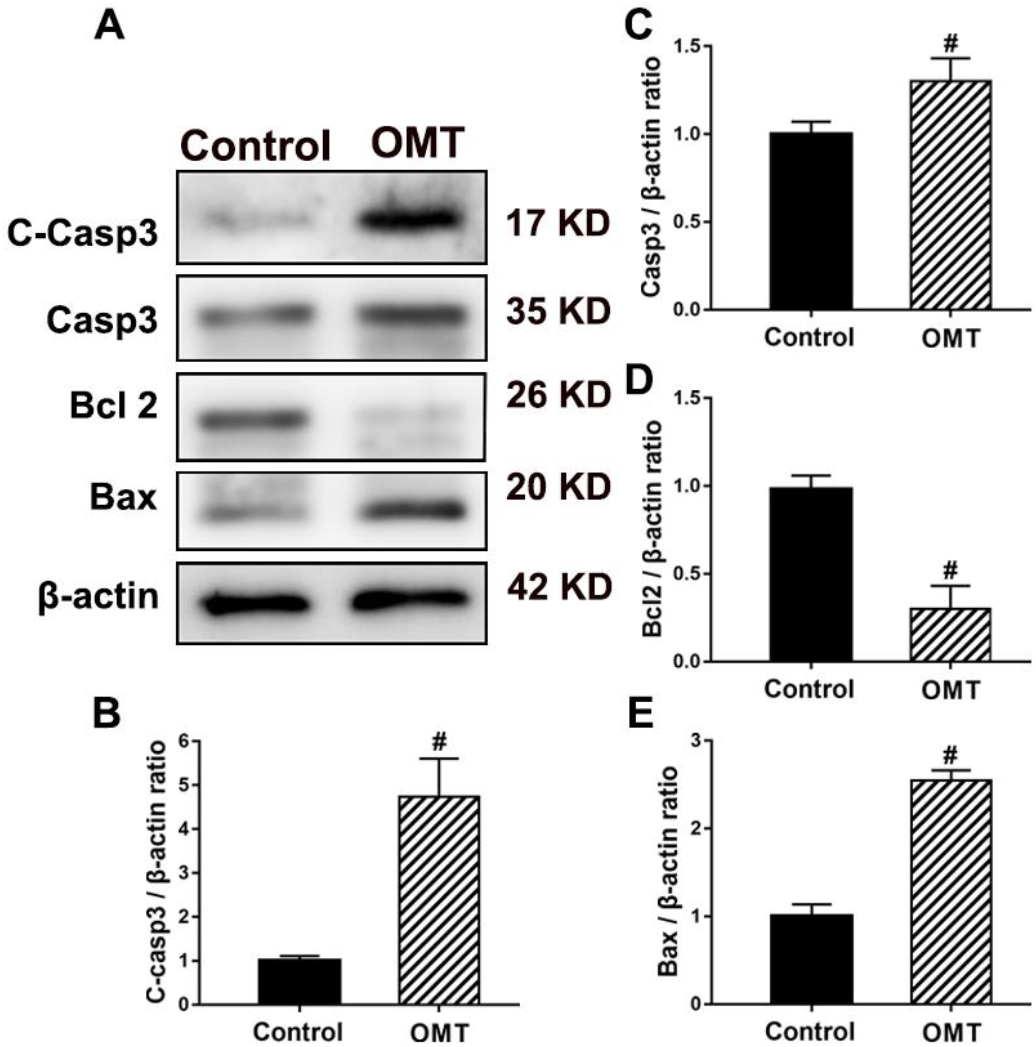
Oxymatrine induces apoptosis of BGC-823 cells (A) Western blot results. (B-E) Western blot quantification histogram.

**Fig. 8.**
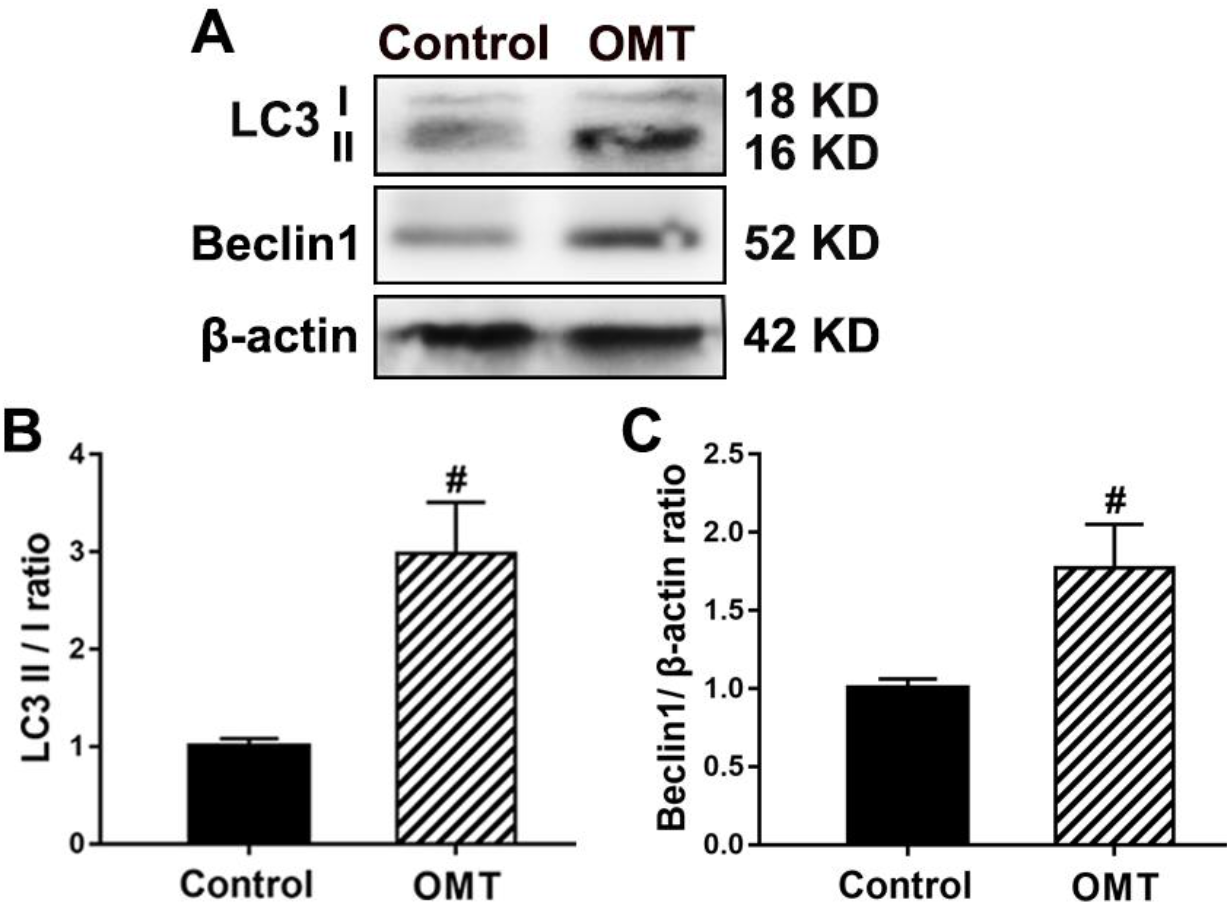
Oxymatrine induces autophagy of BGC-823 cells (A) Western blot results. (B, C) Western blot quantification histogram.

### 2. Expression of apoptosis related proteins in poorly differentiated gastric adenocarcinoma and adjacent tissues

Then we assessed the expression and activation of apoptosis-related proteins including Cleaved caspase3, Caspase3, Bax, and Bcl2, in poorly differentiated gastric adenocarcinoma and adjacent tissues. The western blot results showed that the expression of Cleaved caspase3, Caspase3, Bax in gastric adenocarcinoma is significantly less than that in adjacent tissues. Conversely, the expression of Bcl2 significantly increased in gastric adenocarcinoma compared with adjacent tissues. This result suggested that apoptosis was inhibited in poorly differentiated gastric adenocarcinoma.

### 3. Expression of autophagy-related proteins in poorly differentiated gastric adenocarcinoma and adjacent tissues

Autophagy plays an important role in tumor development and treatment. In addition, we measured the expression of autophagy-related proteins in poorly differentiated gastric adenocarcinoma and adjacent tissues by western blot. The results showed that the ratio of LC3 II/LC3 I in adjacent tissues was significantly higher than that in gastric adenocarcinoma. Similarly, Beclin1 was highly expressed in normal adjacent tissues but dramatically decreased in tumour tissues. This indicated that autophagy was inhibited in poorly differentiated gastric adenocarcinoma.

### 4. Oxymatrine inhibits the growth of BGC-823 cells in vitro

In this experiment, we selected BGC-823 cell lines to detect whether oxymatrine affected the growth and proliferation of poorly differentiated gastric adenocarcinoma cells. We treated the BGC-823 cells with different concentrations of oxymatrine (0, 2, 4, 6, 8 mg/mL) for 24 hours and obtained the cell viability of each group from CCK-8 assay. The results showed that oxymatrine (4, 6, 8 mg/mL) can significantly inhibit the growth of BGC-823 cells. The cell viability gradually decreased with higher matrine concentration in certain concentration-dependent mode. But when the concentration is 6 and 8 mg/mL, the cell inhibition is more serious. Therefore, the concentration of 4 mg/mL was selected for subsequent experiments.

### 5. Oxymatrine inhibits the migration of BGC-823 cells in vitro

Transwell experiment was also conducted to detect the effect of oxymatrine on BGC-823 cells migration. Transwell experiments showed that BGC-823 cells migrated under normal conditions. Compared with the control group, the oxymatrine group significantly reduced the migration of BGC-823 cells after oxymatrine treatment. This result indicated that the oxymatrine inhibited the migration of BGC-823 cells in vitro.

### 6. Oxymatrine inhibits the activation of Slit2/Robo1 signals pathway

In order to verify the effect of oxymatrine on the Slit2/Robo1 signals in BGC-823 cells, western blot was used to detect the key proteins Slit2 and Robo1 in the Slit-Robo signaling pathway. The results showed that Slit2 and Robo1 were highly expressed in BGC-823 cells, but after the intervention of oxymatrine, the expression of Slit2 and Robo1 was significantly reduced. This result suggested that oxymatrine could inhibit the activation of the Slit-Robo signaling pathway in gastric cancer cells.

### 7. Oxymatrine induces apoptosis of BGC-823 cells

In order to investigate the possible molecular mechanisms underlying oxymatrine-induced apoptosis of gastric cancer cells, the expression of Caspase3, Cleaved-caspase3, Bcl2 and Bax was analyzed following treatment with oxymatrine. The results showed that the expression of Caspase 3, Cleaved Capsase 3 and Bax significantly increased in BGC-823 cells following treatment with oxymatrine. On the contrary, the expression of Bcl2 decreased in BGC-823 cells and less after the treatment with oxymatrine. Oxymatrine may therefore reduce gastric cancer cell growth by promoting cell apoptosis in vivo.

### 8. Oxymatrine induces autophagy in BGC-823 cells

Autophagy plays an important role in tumor treatment. On the one hand, autophagy protects tumor cells against drugs; on the other hand, autophagy can promote tumor cell death. Therefore, in order to further study the mechanism of oxymatrine inhibiting the growth of BGC-823 cells, we need to verify whether oxymatrine induces autophagy in BGC-823 cells. Western blot results showed that after the intervention of oxymatrine, the ratio of LC3 II/LC3 I increased significantly. Similarly, the expression of Beclin1 significantly increased in BGC-823 cells following treatment with oxymatrine. This result suggested that oxymatrine induced autophagy in BGC-823 cells.

## Discussion

The present study indicated demonstrates that Slit2 and Robo1 significantly increased in poorly differentiated gastric adenocarcinoma^[9]^. The apoptosis and autophagy are inhibited in poorly differentiated gastric adenocarcinoma^[11]^. OMT inhibits the growth and migration of BGC-823 cells in vitro. The antitumor effects of OMT may be the result of inhibition of cell growth and migration, and inhibits the activation of Slit2/Robo1 signals and induces apoptosis and autophagy. OMT may represent a novel anticancer therapy for the treatment of poorly differentiated gastric adenocarcinoma.

Gastric cancer is one of the most common malignant tumors in humans, and its prognosis is related to the degree of tumor differentiation and metastasis^[1]^. Poorly differentiated gastric adenocarcinoma is a common malignancy of gastric cancer that is more aggressive and has a poor prognosis, mainly due to the biological characteristics of poorly differentiated gastric cancer, which makes it more prone to metastasis. The role of Slit2/Robo1 signals in cancer metastasis remains unclear. Slit2/Robo1 signals have different effects in different situations in cancer cell migration. On the hand, a variety of studies showed that the expression of Slit2 is downregulated or not detected in breast cancer ^[12]^, lung cancer ^[13]^, liver cancer ^[14]^. The inhibitory effect of Slit2/Robo1 is largely related to promoter hypermethylation in these cancers. On the other hand, overexpression of Slit2 and Robo1 appear in gastric cancer ^[15]^, melanoma, pancreatic cancer tissues and hepatocellular carcinoma ^[16]^, which demonstrates that Slit2/Robo1 signals have a facilitating effect in certain cancers. This is consistent with our studies in poorly differentiated gastric adenocarcinoma.

OMT has a variety of pharmacological properties, including anti-inflammatory, anti-oxidative effects, anti-proliferative effects, and anti-apoptotic activities ^[3,4]^. Recent studies have found that OMT has good anticancer activities. OMT has been shown to inhibit tumor cell proliferation in different cancer cells, including breast cancer cell lines (MCF-7), gastric cancer cells (SGC-7901 and MKN45), and human liver cancer cells (SMMC-7721). OMT induces cell cycle arrest and apoptosis via the PI3K/AKT/mTOR pathway in glioma. Preliminary research showed the inhibitory effects of OMT on vascular and lymph node invasion in gastric cancer. This study showed OMT inhibits the growth and migration of BGC-823 cells in vitro. The results showed that Slit2 and Robo1 were highly expressed in BGC-823 cells, but after the intervention of OMT, the expression of Slit2 and Robo1 was significantly reduced. This result suggests that OMT can inhibit the activation of the Slit-Robo signaling pathway in gastric cancer cells.

As we all know, the interaction of autophagy and apoptosis regulates the fate of cells. Autophagy plays an important role in tumor treatment. On the one hand, autophagy protects tumor cells against drugs; on the other hand, autophagy can promote tumor cell death. Apoptosis also plays a central role in cancer therapy. Over-activation of mitochondrial autophagy may degrade essential mitochondria and causes apoptosis. Strikingly, autophagy inhibition or activation might be a therapeutic approach for cancer. Some autophagy-related proteins such as Beclin-1 and LC3 have prognosis for gastric cancer^[17]^. Autophagy and apoptosis are simultaneously involved in deciding the fate of cancer cells, where exists interaction between each other. The present study found that OMT can inhibit the activation of the Slit2/Robo1 signals and induce apoptosis and autophagy in BGC-823 cells at the same time. Previous study showed that the crucial role of Slit2/Robo1 signals in maintaining Lgr5+ stem cell proliferation via regulating autophagy. However, there is still a limitation to this study. It is unknown how Slit2/Robo1 signals mediates the autophagy process and apoptosis.

## Conclusions

Slit2/Robo1 signals significantly increased in poorly differentiated gastric adenocarcinoma. The apoptosis and autophagy are inhibited in poorly differentiated gastric adenocarcinoma. OMT inhibits the growth and migration of BGC-823 cells in vitro. The antitumor effects of OMT may be the result of inhibition of cell growth and migration, and inhibits the activation of Slit2/Robo1 signals and induces apoptosis and autophagy. OMT may represent a novel anticancer therapy for the treatment of poorly differentiated gastric adenocarcinoma.

## Data Availability

All data, models, and code generated or used during the study appear in the submitted article.

## Author Contributions

LQM and MTH conceived and supervised the study; LQM, MTH and TZ designed experiments; TZ, JYH and MTH performed experiments; TZ and MTH analyzed data; TZ and MTH wrote the manuscript; LQM made manuscript revisions. All authors have read and agreed to the published version of the manuscript.

## Compliance with Ethical Standards

### Ethical Approval

The present study was approved by the ethics committee of General Hospital of Ningxia Medical University.

